# Boundary-Specific Failure Modes and Safety Trade-offs of Large Language Models in Chronic Kidney Disease Renoprotective Therapy Review: A Stratified Synthetic Benchmark

**DOI:** 10.64898/2026.05.28.26353938

**Authors:** Shang-En Yeh, Hsuan-Jen Lin, Wu-Wei Lai, Hsuan-Ming Lin

## Abstract

**Background:** Renoprotective therapies - SGLT2 inhibitors, finerenone, and renin-angiotensin system inhibitors (RASi) - remain underutilised in chronic kidney disease (CKD). Large language models (LLMs) may detect therapy omissions, but their performance across CKD severity strata and at clinical decision boundaries has not been evaluated.

**Methods:** We constructed 100 synthetic CKD vignettes (G3a-G5D; 75 with prespecified omissions, 25 decoys) and queried four LLMs three times each at temperature 0 (1,200 calls). Omission criteria were adapted from KDIGO 2024, including an investigator-defined gray-zone RASi initiation criterion at eGFR<15. Two nephrologists independently classified a stratified 20-case subset.

**Results:** For SGLT2 inhibitor and finerenone omissions, all models achieved near-ceiling sensitivity (97-100%). For RASi, performance diverged at the eGFR<15 boundary: Grok 4.1 Fast 85% versus GPT-5.4 55%, Gemini 10%, DeepSeek 10%. Gap-detection inter-rater agreement was perfect (kappa = 1.000). Clinically incorrect reasoning rates ranged from 0% (GPT-5.4) to 27% (DeepSeek R1); of 52 instances, 31 were factual pharmacology errors and 21 reflected conservative boundary-discordant reasoning. Reproducibility (Jaccard) ranged from 0.74 to 0.93.

**Conclusions:** This boundary-aware synthetic benchmark showed that aggregate sensitivity can conceal clinically important operational-rule discordance. Rule-based SGLT2 inhibitor and finerenone omissions were detected with near-ceiling sensitivity, whereas an investigator-defined gray-zone RASi criterion at eGFR<15 exposed model-specific boundary behaviour. Evaluation of LLM-based CKD decision support should report boundary-specific performance, reproducibility, and clinically incorrect reasoning alongside aggregate metrics.

**Key Learning Points:** *What was known:*

- SGLT2 inhibitors, finerenone, and renin-angiotensin system inhibitors reduce kidney and cardiovascular risk in eligible CKD patients but remain underused.
- Large language models are being considered for clinical decision support, but most benchmarks report aggregate accuracy rather than boundary-specific safety.

*This study adds:*

- A boundary-aware synthetic benchmark was constructed to evaluate CKD renoprotective-therapy omission detection across clear rule-based indications, an investigator-defined eGFR<15 gray-zone RASi criterion, decoys, reproducibility, and clinically incorrect reasoning.
- All four evaluated LLMs detected SGLT2 inhibitor and finerenone omissions with near-ceiling sensitivity, but RASi detection diverged sharply at the eGFR<15 boundary, revealing a model-specific conservative non-initiation pattern hidden by aggregate results.
- Safety profiles differed across models: reproducibility ranged from 60% to 89% full agreement, and clinically incorrect reasoning rates ranged from 0% to 27%.

*Potential impact:*

- LLM evaluation for nephrology decision support should report boundary performance, reproducibility, and clinically incorrect reasoning rates alongside aggregate sensitivity.
- Based on the observed boundary discordance and 0–27% clinically incorrect reasoning rate, none of the four evaluated models demonstrated sufficient reproducibility or reasoning accuracy to support unsupervised use for advanced-CKD renoprotective therapy recommendations, particularly around eGFR<15 decisions; comparative human–LLM validation is required before any deployment decision.

## Introduction

Current evaluations of LLM-based clinical decision support rarely test whether models can distinguish clear medication omissions from gray-zone boundary cases where guideline language is sparse. In CKD care, this distinction is central: renoprotective therapies remain substantially underutilised, but inappropriate confidence at advanced-CKD decision boundaries could make aggregate accuracy misleading.

Three drug classes — sodium-glucose cotransporter-2 (SGLT2) inhibitors, finerenone, and renin-angiotensin system inhibitors (RASi) — slow CKD progression in landmark trials [10, 26, 27, 21, 3, 22, 5, 17, 1]. The 2024 KDIGO guideline [24, 16] endorses each with explicit eligibility criteria, yet uptake remains far below indication [29, 33].

Large language models (LLMs) have been proposed as decision-support tools capable of identifying renoprotective therapy gaps [15, 12]. Frontier LLMs perform well on nephrology assessments [32] and generate plausible clinical reasoning [18], yet recent benchmarks reveal important reasoning gaps [28, 23]. Three questions remain before LLMs can be safely positioned for CKD therapy-omission detection. First, no benchmark has evaluated detection across stratified CKD severity, including the eGFR<15 boundary where guideline guidance is least explicit. Second, inter-model variation is underreported; for decision support embedded in clinical workflows, the choice of model has direct safety implications. Third, safety dimensions beyond accuracy - reproducibility across repeated queries [31] and rates of fabricated reasoning [13, 2, 20, 6] - are rarely measured.

This study introduces a boundary-aware synthetic-vignette benchmark for LLM detection of CKD renoprotective therapy omissions. The benchmark separates clear rule-based omissions from investigator-defined eGFR<15 RASi gray-zone cases, embeds decoys to test specificity, repeats queries to quantify reproducibility, and adjudicates clinically incorrect reasoning. This design tests whether aggregate model performance masks clinically relevant boundary discordance.

## Methods

### Study design and ethics

We benchmarked four large language models (LLMs) for detection of renoprotective therapy omissions in chronic kidney disease (CKD), following TRIPOD-LLM reporting guidance [9]. Only investigator-constructed synthetic vignettes were used; no patient data or human subjects were involved. The study was approved by the Institutional Review Board of An Nan Hospital, China Medical University (approval number 113TMANH-REC011(CR-1)). Eligibility rules and gold-standard reasoning were derived from the 2024 KDIGO Clinical Practice Guideline for the Evaluation and Management of Chronic Kidney Disease [24, 16] and contemporary CKD-related trial evidence (DAPA-CKD [10], EMPA-KIDNEY [26], FIDELIO-DKD [3], FIGARO-DKD [22]).

### Case construction

One hundred synthetic case vignettes were constructed and stratified across CKD stages: 30 cases at G3a–G3b (estimated glomerular filtration rate [eGFR] 30–59 mL/min/1.73m^2^), 33 cases at G4 (eGFR 15–29), and 37 cases at G5 / G5D (eGFR <15 or on dialysis). Each vignette specified demographics, eGFR and CKD stage, UACR, serum potassium, blood pressure, HbA1c, comorbidities, current medications, allergies, and a brief clinical question requesting identification of renoprotective gaps. Three omission criteria were adapted from KDIGO 2024, plus an investigator-defined gray-zone RASi rule at eGFR<15:

- **SGLT2 inhibitor** — type 2 diabetes or heart failure with reduced ejection fraction, eGFR ≥20 mL/min/1.73m^2^, UACR ≥200 mg/g, no contraindication recorded (n = 36).
- **Finerenone** — high-risk type 2 diabetic CKD, eGFR ≥25 mL/min/ 1.73m^2^, UACR ≥300 mg/g, serum potassium ≤4.8 mEq/L, on concurrent renin-angiotensin system blockade (n = 34). The primary endpoint was class-level omission detection; background RASi dose optimisation was not a prerequisite, and starting dose was not scored.
- **RASi (ACEi or ARB)** — albuminuric CKD (UACR ≥30 mg/g), serum potassium ≤5.5 mEq/L, no contraindication (n = 35). This included a prespecified eGFR<15 boundary subgroup (n = 20) designed as *de novo* initiation scenarios (no ACEi/ARB in current medications), in which eligibility was operationally preserved when no explicit contraindication was recorded. Initiating RASi at eGFR<15 is not a settled guideline-endorsed recommendation; these cases were included as a deliberate gray-zone benchmark to test conservative heuristics beyond stated criteria. The separate question of whether to *continue* existing RASi at eGFR<15 (STOP-ACEi-relevant) was not tested here.

Thirty cases contained two prespecified gaps each (most commonly SGLT2i+finerenone or SGLT2i+RASi) and 45 contained one gap, yielding 105 prespecified gaps across 75 gap-bearing cases. A further 25 decoy cases without true gaps were constructed using two strategies in combination — current regimens already satisfying all renoprotective indications, and clinical features that operationally precluded one or more candidate classes (e.g., dialysis, documented intolerance) — to assess specificity. The N=100 sample size was prespecified to populate three CKD severity strata (30/33/37) with adequate per-gap counts (36/34/35) for stratified pairwise model comparison while keeping per-case PI annotation time tractable. All vignettes were generated with complete laboratory and medication data by design; no missing values were permitted in the evaluator input. Anthropic Claude Sonnet 4.6 instantiated vignettes and gold-standard reasoning under investigator-defined eligibility rules specified *before* case generation, and was therefore excluded from the evaluator set under a strict generator/evaluator separation.

### Models and evaluation protocol

Four LLMs were evaluated: OpenAI GPT-5.4 (openai/gpt-5.4-20260305), Google Gemini 3.1 Pro (google/gemini-3.1-pro-preview-20260219), xAI Grok 4.1 Fast (x-ai/grok-4.1-fast), and DeepSeek R1 (deepseek/deepseek-r1). All models were accessed on 2026-05-03 through the OpenRouter unified API; identical request interface does not guarantee identical provider infrastructure. Each model received the same structured clinical query, acting as a nephrology clinical pharmacist, to identify any renoprotective gap (drug class, drug, dose, severity, justification) or state explicitly that none existed (full prompt in Supplementary Material). Generation parameters were fixed at temperature 0, max tokens 8192. Each case was queried three times per model (1,200 total LLM calls), enabling intra-model reproducibility assessment. The model-facing prompt instructed models to follow current KDIGO 2024 guidelines and provided simplified eligibility criteria for each drug class; the eGFR<15 boundary rule and specific potassium thresholds used in scoring were not disclosed in the prompt (see Limitations).

### Reproducibility assessment

Free-text responses from the three replicate runs were reduced to the set of issue types identified (SGLT2i, finerenone, RASi, or no-gap), since the clinical unit of interest is the gap type rather than the specific agent. Intra-model consistency was quantified by pairwise Jaccard similarity and full three-way agreement; a consensus run was selected by majority issue-type set (Supplementary Material).

### Outcome classification

All cases were classified against the predefined gold standard by the principal investigator (H-M.L., board-certified nephrologist). A co-investigator nephrologist (H-J.L.) independently classified a stratified 20-case × 4-model sample covering all gap types, the eGFR<15 boundary subgroup, multi-gap cases, and decoy cases. Both raters were blinded to each other but provided the prespecified gold-standard gap labels. Cohen’s κ was computed for three mutually exclusive scoring layers. Gap detection was scored at the issue level (n = 88 ratings), whereas unsupported recommendations and decoy specificity were scored at the model-case level (n = 68 and n = 12, respectively).

An LLM-assisted structured-rubric scorer (GPT-5.4) generated draft TP/FP/FN/TN proposals; both raters made all final classifications, and the PI re-rated 70 ratings under blinded conditions with the draft hidden to confirm no anchoring (Supplementary Material). PI adjudication resolved disagreements.

We additionally recorded *clinically incorrect reasoning* — materially incorrect or unsupported clinical claims in a model’s reasoning text, defined as fabricated guideline thresholds, drug-class properties contradicted by the drug label, or contraindication logic unsupported by trial evidence. Statements matching the study’s prespecified operational criteria (including the UACR ≥300 mg/g finerenone rule) and defensible clinical simplifications were not counted; PI adjudication resolved rater disagreements. Identified instances were stratified post hoc into **factual pharmacology errors** (verifiable against external pharmacology or label text, independent of our operational criteria) and **boundary-discordant conservative reasoning** (a more restrictive potassium or eGFR threshold than our criterion, possibly reflecting defensible conservatism). The former is the headline safety finding; the latter is reported separately.

### Statistical analysis

Per-omission-type sensitivity and overall specificity were estimated with Wilson 95% confidence intervals [30]. Overall precision was computed across all FPs in the cohort. Overall F1 was estimated by case-level non-parametric bootstrap (n = 2,000 resamples; random seed = 42 for reproducibility). Inter-model differences in detection of each omission type were tested by Cochran’s Q test [7]; if omnibus p < 0.05, all six pairwise model comparisons were performed by McNemar’s test [19] and adjusted within each gap type by the Holm step-down procedure [11]. Effect sizes were reported as Cohen’s h. Inter-rater reliability between the two nephrologist raters was reported as Cohen’s κ [8] with the Landis–Koch interpretation framework [14].

Two prespecified secondary analyses were conducted: RASi sensitivity in the eGFR<15 boundary subgroup versus eGFR≥15, and intra-rater κ between the PI’s anchored and blinded passes (Supplementary Material). Clinically incorrect reasoning rate was analysed as an exploratory secondary safety endpoint with case-level proportions and Wilson 95% CIs; formal inferential testing was not performed (case-paired endpoint precluding standard two-sample tests). This analysis is hypothesis-generating and requires real-world replication.

All analyses were performed in Python 3.13 with NumPy 2.4, SciPy 1.17, and Matplotlib 3.10. Reproducibility materials are described in the Data Availability statement.

## Results

### Cohort

The 100 case vignettes (Table 1) had a median eGFR of 24 mL/min/1.73m^2^ (IQR 11–36), with stratified representation across G3a–G3b (n = 30), G4 (n = 33), and G5/G5D (n = 37); type 2 diabetes was present in 78 cases and hypertension in 100. The 75 gap-bearing cases contained 105 prespecified gaps (36 SGLT2 inhibitor, 34 finerenone, 35 RASi), with 30 multi-gap cases; 25 decoy cases were included to assess specificity. Per-gap recall in multi-gap cases was 100% for three models and 98.3% for DeepSeek R1 (Supplementary Table S1).

**Table 1.** Case Characteristics (n=100 synthetic CKD vignettes)

| Characteristic | Overall (n=100) | GA — G3a/G3b (n=30) | GB — G4 (n=33) | GC — G5/G5D (n=37) |
| --- | --- | --- | --- | --- |
| **Demographics** |  |  |  |  |
| Age, years, median (IQR) | 68 (58–74) | 69 (59–77) | 65 (58–74) | 68 (60–74) |
| Male sex, n (%) | 67 (67%) | 20 (67%) | 22 (67%) | 25 (68%) |
| BMI, kg/m², median (IQR) | 26 (23–30) | 27 (23–30) | 26 (22–28) | 27 (25–30) |
| **Renal function & labs** |  |  |  |  |
| eGFR, mL/min/1.73m², median (IQR) | 24 (11–36) | 48 (41–53) | 25 (21–27) | 9 (6–12) |
| UACR, mg/g, median (IQR) | 270 (118–358) | 350 (285–420) | 350 (280–400) | 100 (70–170) |
| Serum potassium, mEq/L, median (IQR) | 5 (4–5) | 4 (4–5) | 5 (4–5) | 5 (5–5) |
| HbA1c, %, median (IQR) | 8 (7–8) | 8 (7–8) | 8 (7–8) | 8 (7–8) |
| Systolic BP, mmHg, median (IQR) | 149 (141–156) | 139 (132–146) | 150 (145–156) | 155 (150–166) |
| **Comorbidities, n (%)** |  |  |  |  |
| Type 2 diabetes mellitus | 78 (78%) | 27 (90%) | 28 (85%) | 23 (62%) |
| Hypertension | 100 (100%) | 30 (100%) | 33 (100%) | 37 (100%) |
| Heart failure | 5 (5%) | 3 (10%) | 2 (6%) | 0 (0%) |
| Dyslipidemia | 19 (19%) | 10 (33%) | 6 (18%) | 3 (8%) |
| On dialysis, n (%) | 20 (20%) | 0 (0%) | 0 (0%) | 20 (54%) |
| Medications per case, median (IQR) | 10 (8–11) | 8 (8–8) | 10 (10–10) | 11 (11–11) |

Gap Type Distribution (gold standard, non-decoy cases)
| Gap type | n cases | % of gap cases |
| --- | --- | --- |
| SGLT2 inhibitor | 36 | 48% |
| Finerenone | 34 | 45% |
| RASi (ACEi/ARB) | 35 | 47% |
| Decoy cases (no true gap) | 25 | — |

Notes
- All values are descriptive; cases were constructed to satisfy KDIGO 2024 eligibility criteria for the assigned gap type. - Obesity diagnoses, where present, were assigned based on physician documentation rather than BMI alone (e.g., metabolic obesity in normal-weight individuals); discordance between BMI and obesity diagnosis in 3 cases was retained for clinical realism. - A case may contain multiple gap types (e.g., G1+G2); gap counts above sum to total gaps, not unique cases.

### Overall performance differs by drug class and model

Across all 100 cases, the four LLMs differed substantially in overall classification performance (Table 2). Sensitivity ranged from 97.1% (Grok 4.1 Fast, 95% CI 92–99%) to 80.0% (DeepSeek R1, 71–87%), with GPT-5.4 91.4% (85–95%) and Gemini 3.1 Pro 81.9% (73–88%); F1 followed the same ranking (Grok 0.981, GPT-5.4 0.932, Gemini 0.887, DeepSeek 0.853). Decoy specificity was 100.0% (Gemini), 96.0% (Grok), 92.0% (GPT-5.4), and 88.0% (DeepSeek) (Figure 4).

**Table 2.**
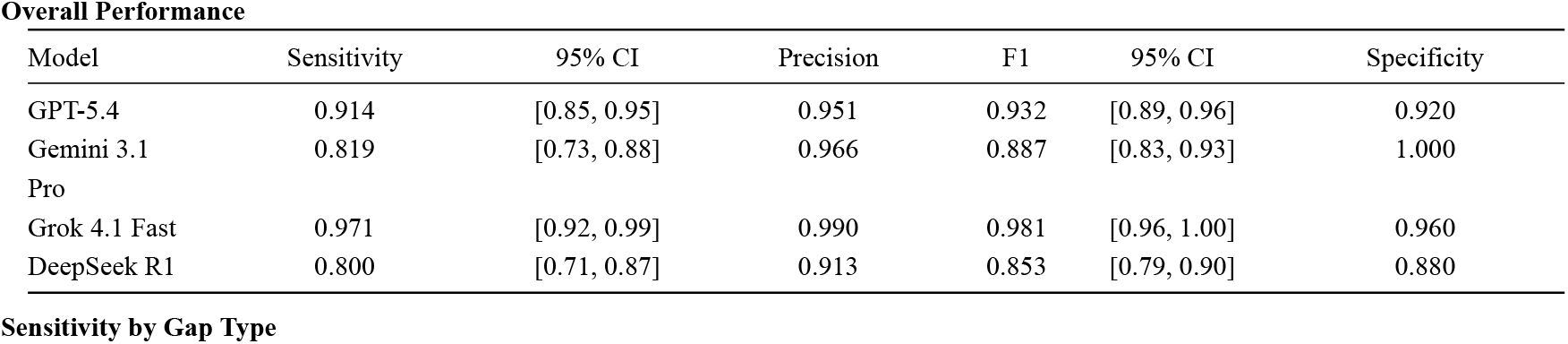
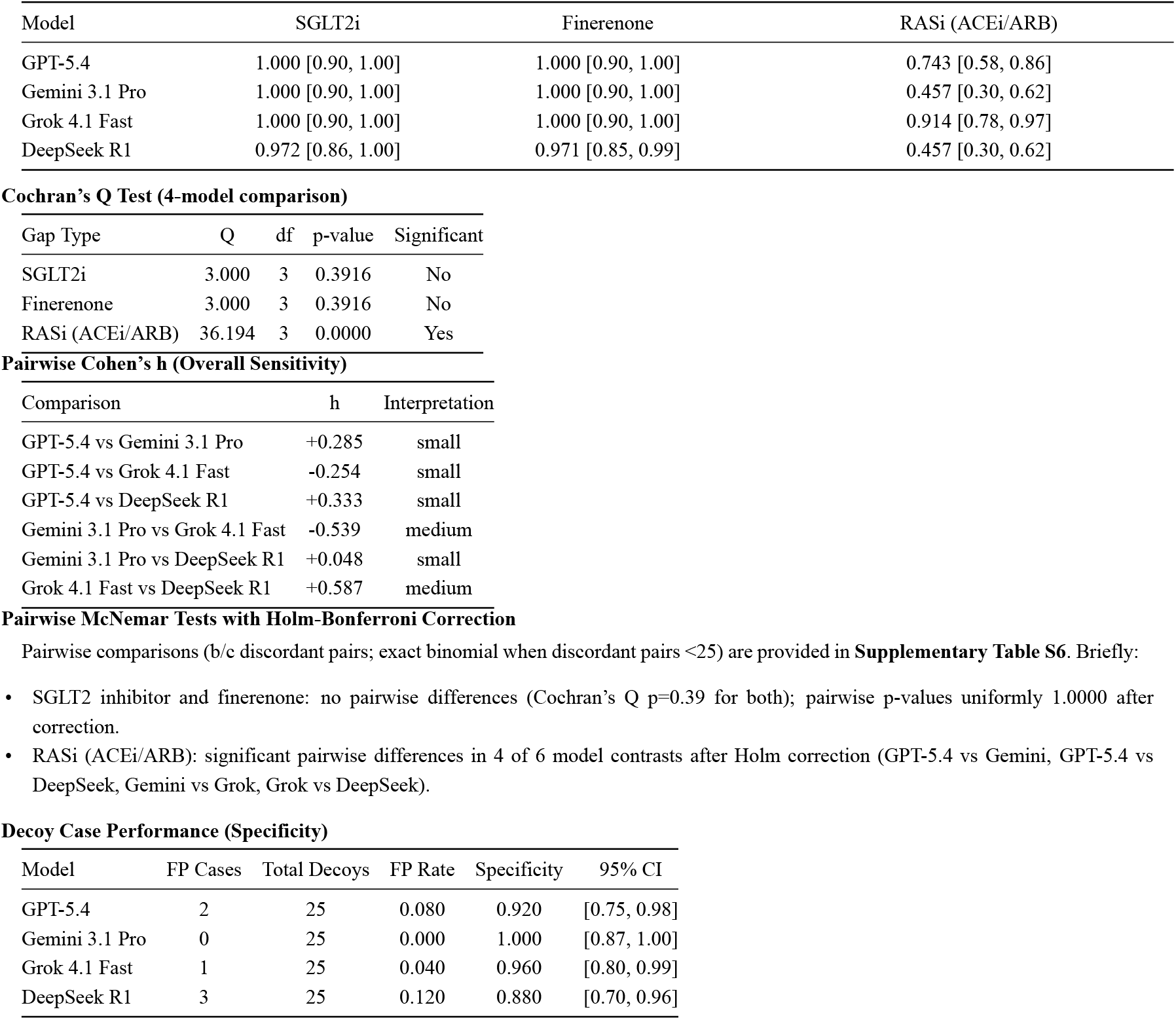
Overall Performance and Statistical Tests.

### Rule-based omissions show near-ceiling detection

Sensitivity for SGLT2 inhibitor and finerenone omissions was near-perfect across all four models: 100% (95% CI 90.4–100.0%) for GPT-5.4, Gemini 3.1 Pro, and Grok 4.1 Fast on both classes, and 97.2% (95% CI 85.8–99.5%) for SGLT2 inhibitor and 97.1% (95% CI 85.1– 99.5%) for finerenone for DeepSeek R1 (Figure 2). Cochran’s Q detected no significant inter-model difference for these two omission types (SGLT2i Q = 3.00, df = 3, p = 0.39; finerenone Q = 3.00, p = 0.39).

**Figure 1.**
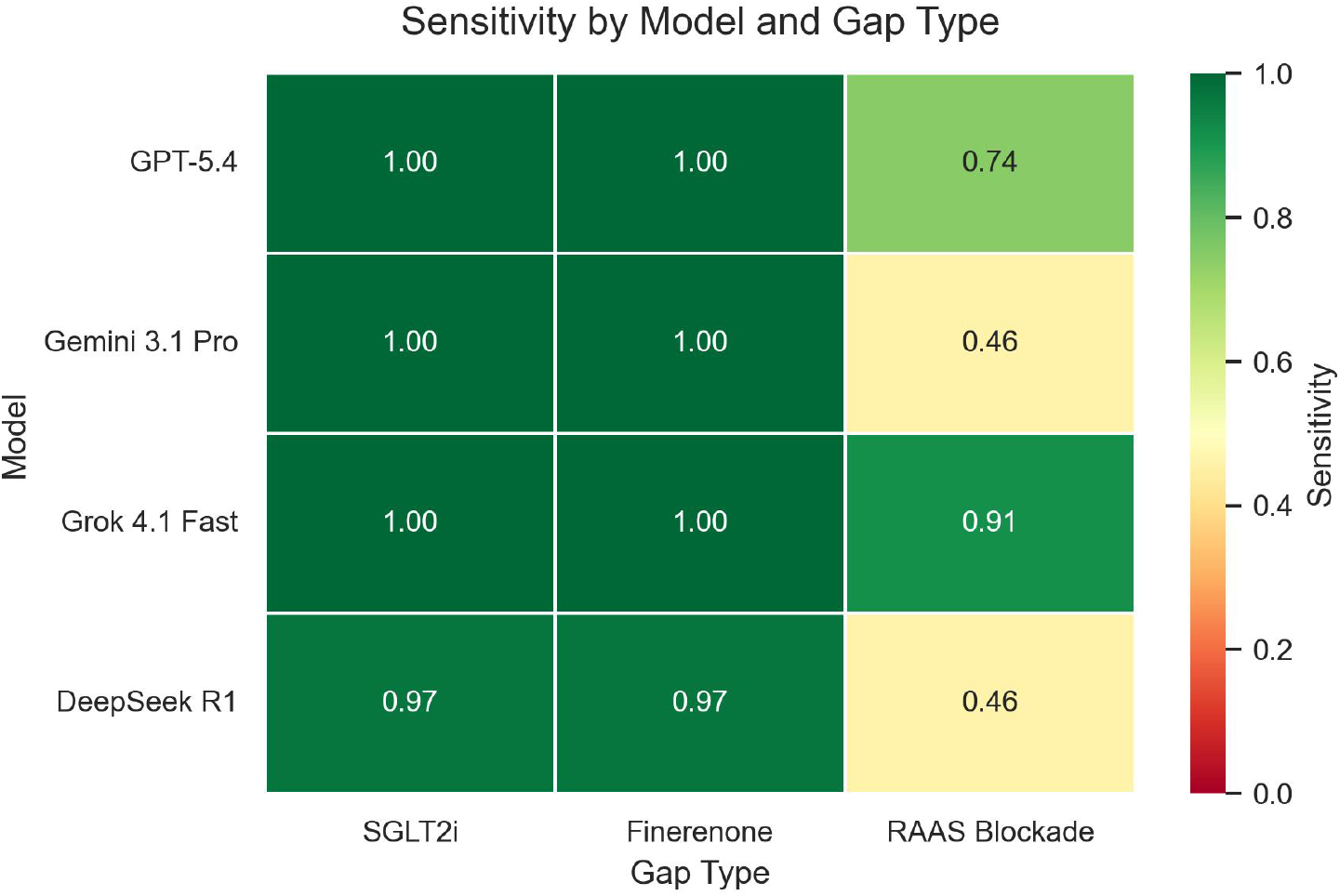
Overall Performance - Sensitivity by Model and Gap Type. Heatmap of per-omission-type sensitivity across the four large language models evaluated. Each cell shows the proportion of prespecified gaps of that omission type correctly identified. SGLT2 inhibitor and finerenone omissions were detected at near-ceiling sensitivity by all four models; RASi detection diverged substantially between models, driven primarily by the eGFR<15 boundary subgroup (see Figure 3). Colour gradient: red (low), yellow (intermediate), green (high). Alt text: Heatmap showing four models by three drug classes; SGLT2i and finerenone cells are green (high sensitivity) while RASi cells vary from green to red.

**Figure 2.**
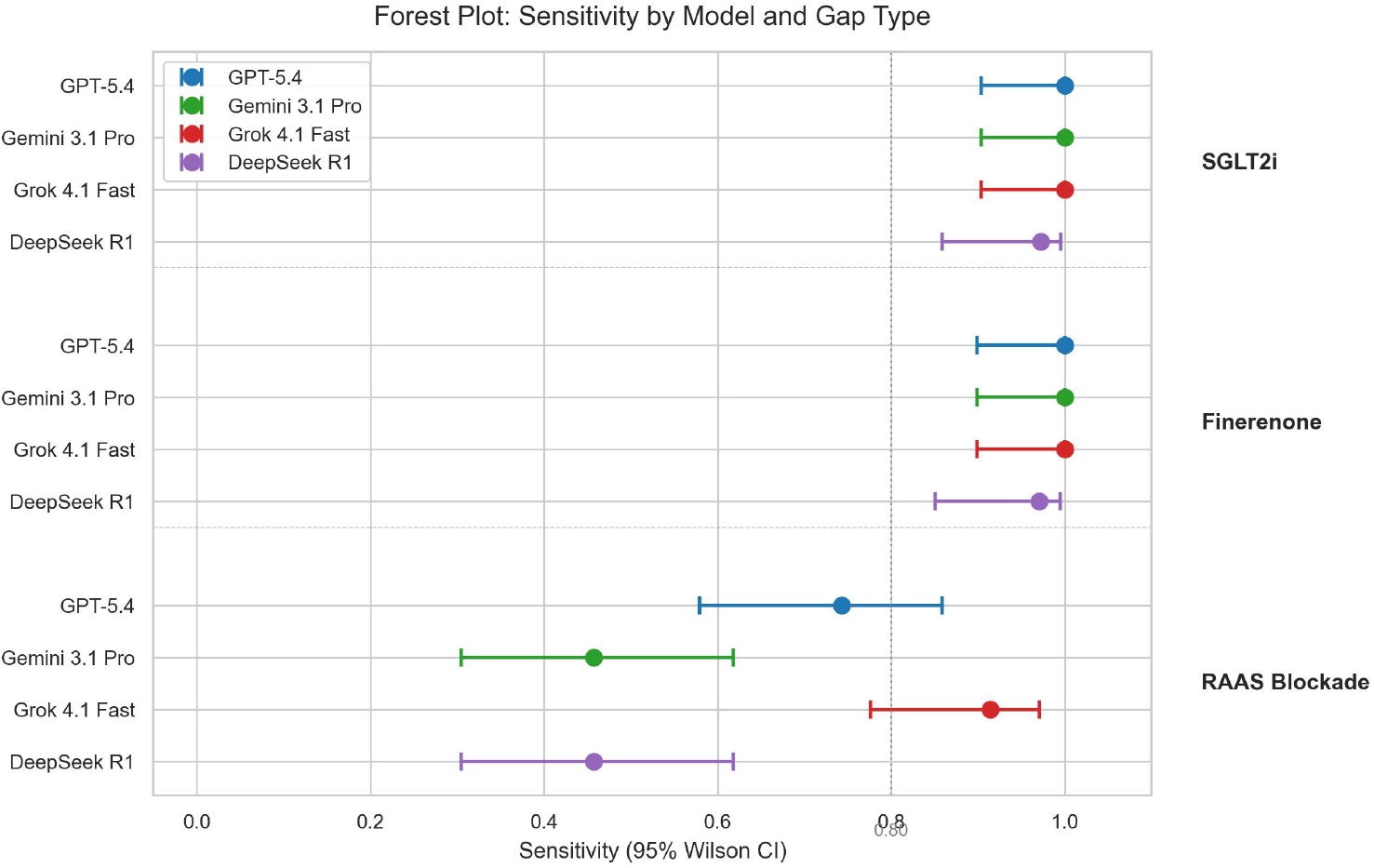
Sensitivity Forest Plot with 95% Wilson Confidence Intervals. Per-model, per-omission-type sensitivity point estimates with Wilson 95% confidence intervals. The vertical reference line at 0.80 marks a conventional benchmark for clinically usable sensitivity. RASi detection shows the widest inter-model spread; SGLT2 inhibitor and finerenone detection cluster near the ceiling. Alt text: Forest plot with point estimates and horizontal confidence interval bars for each model-drug combination; RASi shows the widest inter-model spread.

In contrast, RASi detection diverged markedly between models: sensitivity 91.4% (95% CI 77.6–97.0%) for Grok 4.1 Fast, 74.3% (95% CI 57.9–85.8%) for GPT-5.4, 45.7% (95% CI 30.5–61.8%) for both Gemini 3.1 Pro and DeepSeek R1; Cochran’s Q = 36.19, df = 3, p < 0.001. Holm-corrected pairwise McNemar tests confirmed significant differences between Grok 4.1 Fast and Gemini 3.1 Pro (p = 0.0002), Grok 4.1 Fast and DeepSeek R1 (p = 0.0002), GPT-5.4 and DeepSeek R1 (p = 0.008), and GPT-5.4 and Gemini 3.1 Pro (p = 0.019). The Gemini 3.1 Pro versus DeepSeek R1 comparison (1 discordant pair in each direction; p = 1.000) and the Grok 4.1 Fast versus GPT-5.4 comparison (Grok detected 6 cases that GPT-5.4 missed and zero in the reverse direction; p = 0.063) did not meet significance after correction.

### eGFR<15 boundary subgroup

The inter-model gap in RASi detection was localised to the prespecified eGFR<15 boundary subgroup (Figure 3). Among 20 boundary cases — operationally defined as RASi-eligible under the study’s gray-zone criterion (UACR ≥30, K ≤5.5, no contraindication) — sensitivity was 85% (17/20) for Grok 4.1 Fast, 55% (11/20) for GPT-5.4, 10% (2/20) for Gemini 3.1 Pro, and 10% (2/20) for DeepSeek R1. Among 15 non-boundary RASi cases (eGFR≥15), sensitivity was uniformly high: 100% for GPT-5.4 and Grok 4.1 Fast, 93% for both DeepSeek R1 and Gemini 3.1 Pro. The boundary– non-boundary sensitivity gap was −15 (Grok), −45 (GPT-5.4), −83 (DeepSeek), and −83 (Gemini) percentage points, identifying eGFR<15 as the principal locus of conservative RASi non-initiation (Table 3).

**Figure 3.**
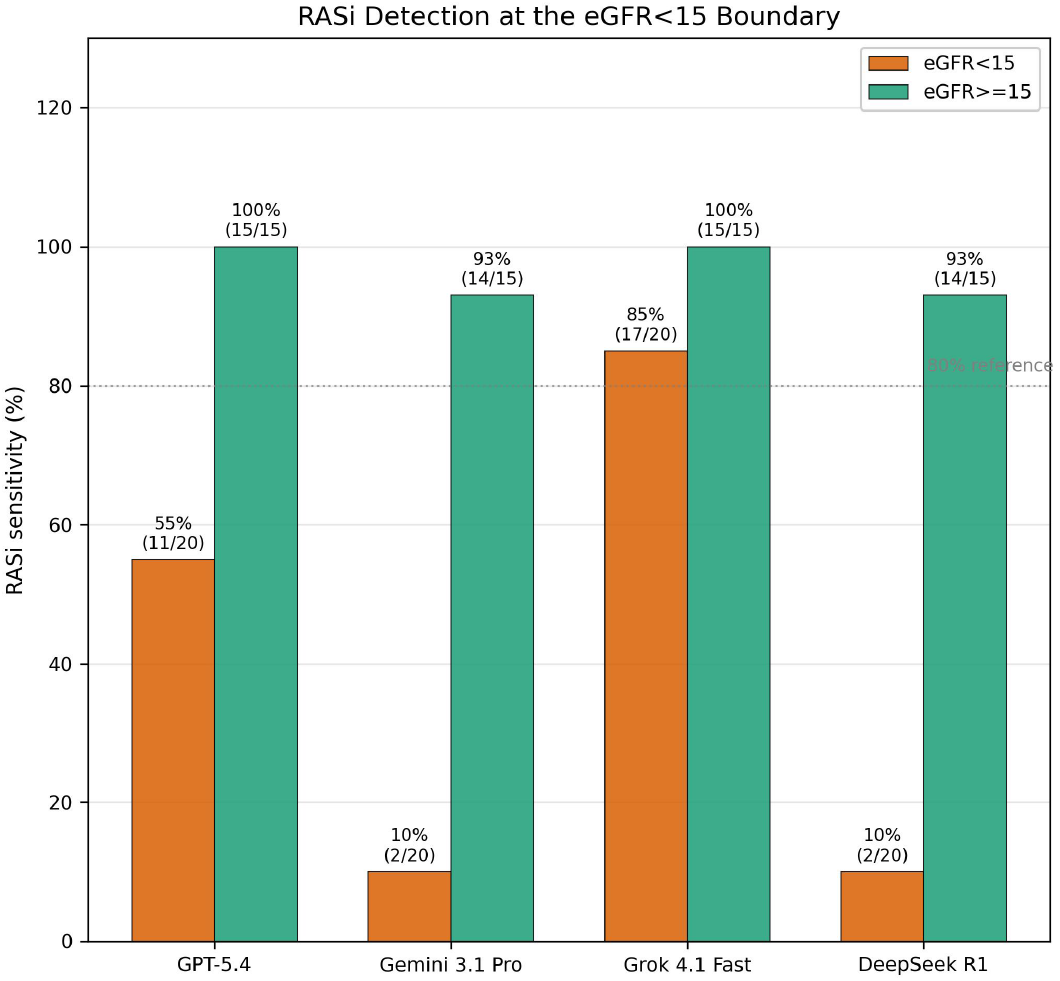
eGFR<15 Boundary Subgroup - RASi Detection. Sensitivity of RASi detection in the prespecified eGFR<15 boundary subgroup (n = 20) versus non-boundary RASi cases (eGFR>=15, n = 15). All four models maintained sensitivity at or near 100% in non-boundary cases; in the boundary subgroup, only Grok 4.1 Fast preserved high sensitivity (85%), while GPT-5.4 dropped to 55% and both Gemini 3.1 Pro and DeepSeek R1 dropped to 10%. The boundary-minus-non-boundary difference localises a conservative-bias regime activated below the eGFR threshold at which guideline guidance is sparsest. Alt text: Grouped bar chart comparing RASi sensitivity at eGFR below 15 versus eGFR 15 and above; Grok maintains high sensitivity at the boundary while three other models drop sharply.

**Figure 4.**
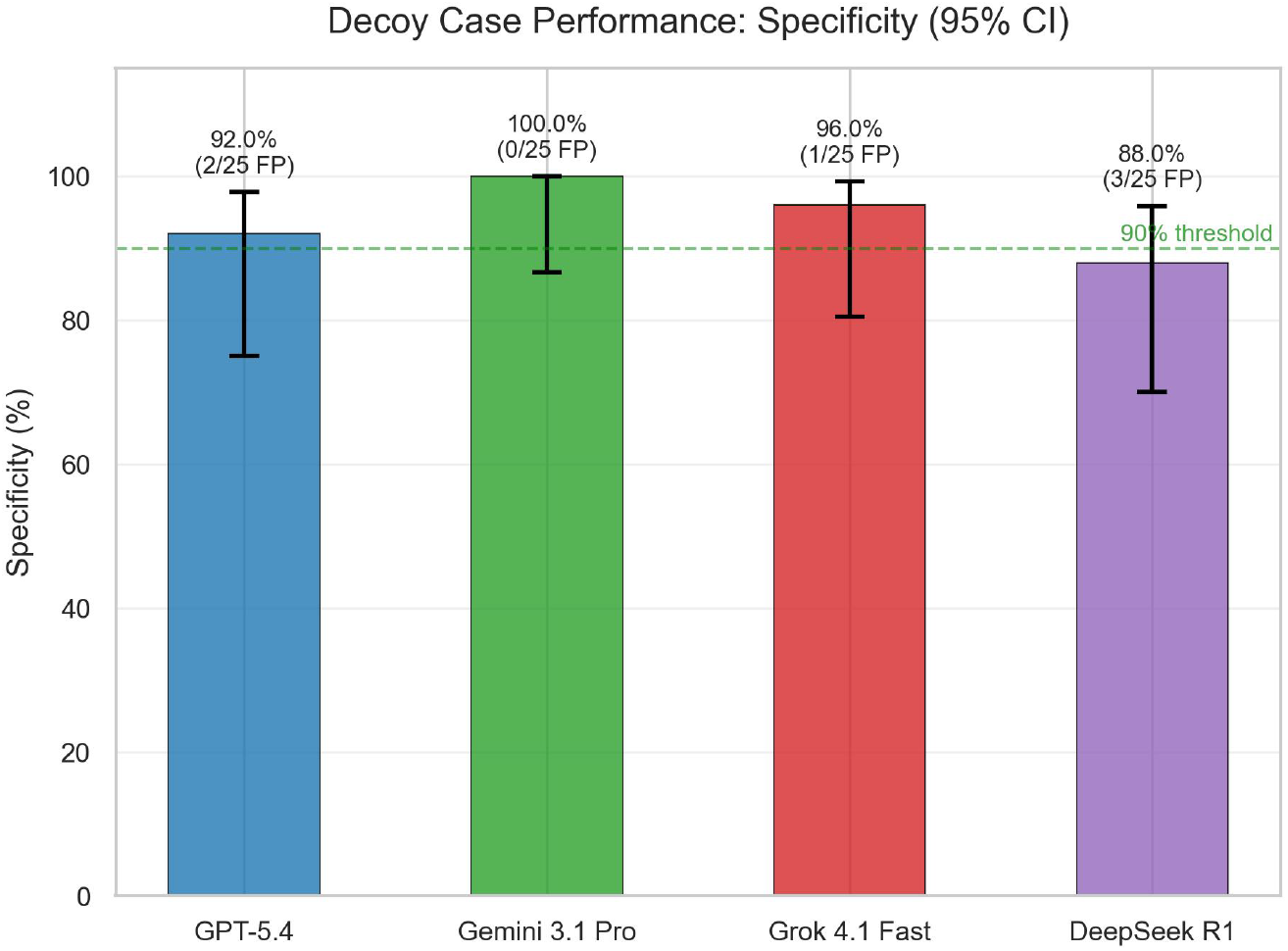
Decoy Specificity - False-Positive Rate on No-Gap Cases. Specificity, defined as the proportion of decoy cases correctly identified as having no true treatment gap, with Wilson 95% confidence intervals across the 25 decoy cases. The horizontal reference line marks 90% specificity. Specificity ranged from 88% to 100%; three models met or exceeded the 90% reference line, while DeepSeek R1 was 88%. False-positive cases were dominated by extra recommendations of losartan titration or finerenone addition in cases without the prespecified gap. Alt text: Bar chart of decoy specificity with confidence intervals for four models; all exceed 88 percent, Gemini reaches 100 percent.

**Table 3.** Error Taxonomy (False Negatives by Stratum, Model, and Drug Class)

False Negatives by Gap Type and eGFR Stratum
| Gap type | eGFR<15 | eGFR 15–29 | eGFR 30–59 | Total |
| --- | --- | --- | --- | --- |
| SGLT2 inhibitor | 0 | 1 | 0 | 1 |
| Finerenone | 0 | 0 | 1 | 1 |
| RASi (ACEi/ARB) | 48 | 2 | 0 | 50 |

False Negatives by Model and Gap Type
| Model | SGLT2 inhibitor | Finerenone | RASi | Total |
| --- | --- | --- | --- | --- |
| GPT-5.4 | 0 | 0 | 9 | 9 |
| Gemini 3.1 Pro | 0 | 0 | 19 | 19 |
| Grok 4.1 Fast | 0 | 0 | 3 | 3 |
| DeepSeek R1 | 1 | 1 | 19 | 21 |

False Positives by Source (decoy vs non-decoy) and Drug Class
| Model | FP in decoys (no gap) | FP in gap cases (extra rec) | Total |
| --- | --- | --- | --- |
| GPT-5.4 | 2 | 3 | 5 |
| Gemini 3.1 Pro | 0 | 3 | 3 |
| Grok 4.1 Fast | 1 | 0 | 1 |
| DeepSeek R1 | 3 | 5 | 8 |

### Robustness and extension of the boundary finding

Two prespecified supplementary analyses tested robustness of the eGFR<15 finding and a second clinical boundary (Supplementary Tables S3 and S4). The conservative bias was graded rather than threshold-defined: as the cutoff widened from eGFR<10 to <25, RASi sensitivity rose monotonically for the three lower-recall models (e.g., GPT-5.4 0/9 → 18/27), while Grok remained ≥67%. Finerenone detection was unaffected by potassium stratum (100% in K 4.6–4.8). RASi detection in the K 5.1–5.5 yellow zone (n = 7) dropped further for Gemini and DeepSeek (14% each); stratum size precludes formal inference. The conservative bias is therefore graded with respect to eGFR but selective with respect to drug class and potassium.

### Repeated queries reveal model-specific reproducibility

Across the three replicate queries per case, intra-model agreement on the identified issue-type set differed substantially between models (Supplementary Material). Full agreement across all three runs occurred in 89% of cases for Grok 4.1 Fast, 82% for GPT-5.4, 81% for Gemini 3.1 Pro, and 60% for DeepSeek R1. Mean pairwise Jaccard similarity was 0.93 (Grok), 0.88 (GPT-5.4), 0.87 (Gemini), and 0.74 (DeepSeek). Five DeepSeek R1 cases (5%) showed three-way disagreement; no other model produced any three-way discordant case. All primary analyses used a single consensus decision per model–case.

### Inter-rater agreement and methodological verification

On the 20-case inter-rater subset, gap detection agreement was perfect (κ = 1.000, 88/88). For unsupported recommendations, observed agreement was 97.1% (66/68); the near-absence of positive events precluded meaningful κ (prevalence-dependent paradox). All 12 decoy-model pairs were unanimously true negatives. The PI’s blinded re-rating yielded intra-rater κ = 0.971 vs the draft-anchored pass, confirming no draft anchoring (Supplementary Material).

### Clinically incorrect reasoning (exploratory secondary endpoint)

Across 100 cases, 52 instances of clinically incorrect claims were identified: 31 (60%) were **external factual pharmacology errors** independent of the study’s operational criteria (e.g., fabricated sulfonamide cross-reactivity, unsupported ARB-over-ACEi hyperkalemia claims, inverted sodium bicarbonate pharmacology), and 21 (40%) were **conservative boundary reasoning** applying stricter potassium or eGFR thresholds than the operational criterion. The case-level rate (either category) differed markedly across models (Figure 5): 0/100 (0.0%, 95% CI 0.0–3.7%) for GPT-5.4, 10/100 (10.0%) for Grok 4.1 Fast, 9/100 (9.0%) for Gemini 3.1 Pro, and 27/100 (27.0%, 19.3–36.4%) for DeepSeek R1. Non-overlapping Wilson CIs between GPT-5.4 and all other models suggest meaningful between-model differences, but formal pairwise inferential testing was not performed for this exploratory case-paired endpoint; findings require real-world replication.

**Figure 5.**
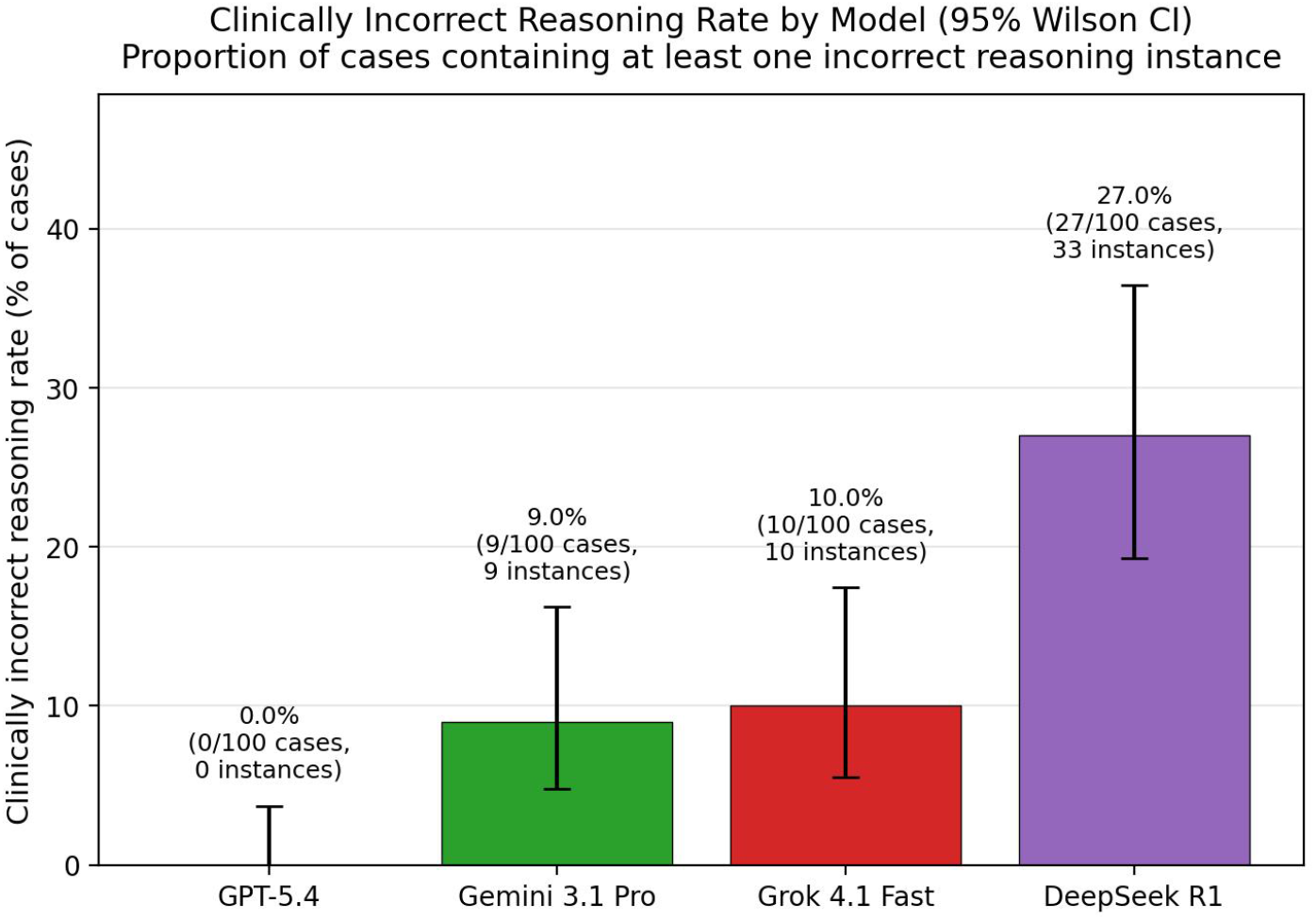
Clinically Incorrect Reasoning Rate by Model - Exploratory Safety Endpoint. Case-level clinically incorrect reasoning rate, defined as the proportion of cases containing at least one factual pharmacology error or boundary-discordant conservative reasoning claim in the model reasoning text, with Wilson 95% confidence intervals. GPT-5.4 produced no clinically incorrect reasoning across 100 cases; DeepSeek R1 produced clinically incorrect reasoning in 27% of cases. Identified instances concerned clinical pharmacology, including fabricated guideline thresholds, incorrect drug-class properties, and flawed contraindication logic. This analysis is positioned as a secondary, exploratory safety endpoint; pairwise rate differences are descriptive, and no formal inferential testing was performed. Alt text: Bar chart of case-level clinically incorrect reasoning rates; GPT-5.4 at zero percent, DeepSeek at 27 percent, with Wilson confidence interval error bars.

## Discussion

Among four LLMs evaluated for renoprotective therapy omission detection, three diverged substantially from an investigator-defined operational criterion at the eGFR<15 RASi boundary, while all performed near-ceiling on SGLT2 inhibitor and finerenone omissions. Reproducibility varied markedly (full-agreement 60-89%), and clinically incorrect reasoning rates spanned a 27-percentage-point range (0–27%).

### Rule-based versus boundary detection

For SGLT2 inhibitor and finerenone omissions, all models achieved 97-100% sensitivity with no inter-model difference (Cochran’s Q p = 0.39), reflecting unambiguous eligibility logic. For RASi, overall sensitivity ranged from 46% to 91% (Q < 0.001), localised to the eGFR<15 boundary subgroup: sensitivity was 85% for Grok 4.1 Fast, 55% for GPT-5.4, and 10% for both Gemini and DeepSeek. The supplementary threshold analysis confirmed that this is graded, not threshold-defined: GPT-5.4 and DeepSeek reached 0% at eGFR<10. The STOP-ACEi trial [4] demonstrated safety of *continuing* existing RASi at eGFR<30, but all 20 boundary cases tested *de novo initiation* (no ACEi/ARB prescribed). Models did not violate trial evidence; they diverged from a prespecified operational gray-zone criterion extending initiation into a zone without dedicated randomised evidence. Characterising this selective operational-rule discordance is important because decision-support systems must behave predictably when local rules extend beyond explicit trial evidence.

Potassium-stratum analysis showed no conservative behaviour for finerenone at borderline K (4.6–4.8: 100% detection) yet further RASi reductions at K 5.1–5.5, consistent with drug-eligibility-specific rather than universal boundary conservatism [23].

### Reproducibility and clinically incorrect reasoning

Full agreement across three replicate queries ranged from 89% (Grok) to 60% (DeepSeek), with five DeepSeek cases showing three-way disagreement. This differential has direct deployment implications: a model whose classification varies across identical queries presents a barrier to quality-improvement workflows requiring reproducible recommendations.

GPT-5.4 produced zero clinically incorrect reasoning instances (0.0%, CI 0.0–3.7%) while DeepSeek produced 27%. Such instances were more common in gap-bearing than decoy cases, suggesting that clinically incorrect reasoning is most likely at the moment a recommendation is being constructed rather than during null-case review.

### No single model dominates

The four models occupied distinct positions: Grok 4.1 Fast led on sensitivity (97%) and reproducibility (89%) but produced clinically incorrect reasoning in 10% of cases, while GPT-5.4 produced none (0% vs 9–27% in the other three models) but underperformed at the boundary (55%). Relative performance in real clinical documentation, and relative to nephrologist reasoning, was not measured here and remains to be established. Within this benchmark, no model provided sufficient boundary stability to support unsupervised use for eGFR<15 RASi decisions.

### Boundary-aware reporting is required before deployment claims

Aggregate sensitivity may overstate clinical readiness; boundary-specific performance, reproducibility, and clinically incorrect reasoning rates should accompany aggregate metrics [25]. Until these dimensions are systematically validated, synthetic-benchmark results should not be interpreted as clinical-readiness evidence for LLM-assisted renoprotective therapy review. Prospective validation against real-world clinical documentation is required and is planned as a separate study.

### Limitations

First, **synthetic English-language vignettes do not capture real clinical narratives** - inconsistent documentation, missing values, non-English contexts, or patient-reported barriers. A real-world replication is planned pending institutional review board approval.

Second, **inter-rater reliability was assessed on a stratified 20-case subset**, not the full cohort. The decoy subset (n = 12) is particularly small for stable kappa estimation. Cases outside the subset were classified by the principal investigator only.

Third, **only four LLMs were tested at a single time point using a single prompt strategy at temperature 0**. The model landscape changes rapidly; findings may not generalise to other systems, prompt formulations, or temperature settings.

Fourth, **the eGFR<15 RASi criterion was investigator-defined and intentionally tested a gray-zone de novo initiation scenario**, not a settled guideline mandate or continuation question. Because the prompt did not disclose this scoring boundary, boundary misses should be interpreted as operational-rule discordance rather than proven clinical-reasoning failure. The clinically incorrect reasoning analysis was exploratory with descriptive reporting only.

Fifth, **Claude Sonnet 4.6 was used as case generator** and excluded from evaluation; this reduces but does not eliminate generator-evaluator bias. Inter-rater agreement measures scoring consistency against the prespecified gold standard, not independent gold-standard derivation.

## Conflicts of Interest

The authors declare no conflicts of interest.

## Funding

No external funding was received.

## Data Availability

The complete reproducibility package underlying this study — 100 synthetic case vignettes, gold-standard definitions, scoring rubrics, the full evaluation prompt, all 1,200 model run outputs, principal-investigator and co-rater scored files, and analysis scripts — is available immediately upon request to the corresponding author, including review-stage access for the handling editor and peer reviewers. No patient-level data were used. A versioned public repository will be deposited upon acceptance.

## Author Contributions

H-M.L. designed the study, constructed cases and gold standards, performed primary classification, and conducted analysis. S-E.Y. drafted and revised the manuscript and reviewed the analytical data. H-J.L. independently classified the inter-rater reliability subset. W-W.L. contributed to manuscript drafting and revision. All authors reviewed and approved the final manuscript.

## Acknowledgements

The authors thank the model and API providers and the OpenRouter platform for API access. No non-author human writing or analytical assistance was used. Generative AI roles in case construction, response extraction, and draft scoring are disclosed in Methods. This study was approved by the Institutional Review Board of An Nan Hospital, China Medical University (approval number 113TMANH-REC011(CR-1)).

## Notes

### Competing Interest Statement

The authors have declared no competing interest.

### Author Declarations

This study wasapproved by the Institutional Review Board of An Nan Hospital, China Medical University (approval number 113TMANH-REC011(CR-1))

