## Supplementary for "Boundary-Specific Failure Modes and Safety Trade-offs of Large Language Models in Chronic Kidney Disease Renoprotective Therapy Review: A Stratified Synthetic Benchmark"

#### Contents


**Supplementary Material S0: Full LLM Evaluation Prompt** **1**

**Supplementary Table S1: Multi-Gap Case Performance** **2**

**Supplementary Table S2: Clinically Incorrect Reasoning Analysis** **3**

**Supplementary Table S3: eGFR Threshold Robustness** **7**

**Supplementary Table S4: Potassium Boundary Subgroup** **8**

**Supplementary Table S5: Replicate-Query Sensitivity** **9**

**Supplementary Table S6: Pairwise McNemar Tests** **10**


ewpage

#### Supplementary Material S0: Full LLM Evaluation Prompt

##### System Prompt

> You are a nephrology clinical pharmacist specializing in chronic kidney disease (CKD) management. Your task is to review a CKD patient's current medication list and identify renoprotective therapy OMISSIONS — medications that are indicated by current KDIGO 2024 guidelines but are NOT currently prescribed.
>
> Focus specifically on these three drug classes:
>
> 1. SGLT2 inhibitors (e.g., dapagliflozin, empagliflozin) — indicated for T2DM + CKD with significant albuminuria (UACR ≥200 mg/g) or HFrEF, when eGFR ≥20.
> 2. Finerenone — indicated for T2DM + CKD + UACR ≥300 mg/g + eGFR ≥25 + K ≤4.8, when already on RASi (ACEi/ARB).
> 3. RASi (ACEi or ARB) — indicated for CKD with albuminuria (UACR ≥30 mg/g).
>
> For each identified gap:
>
> - State the drug class that is missing.
> - Explain why it is indicated for this patient (cite specific clinical parameters from the case).
> - Recommend a specific medication with starting dose.
> - Assess severity: Critical (delays proven renoprotection), Major (important but less urgent), or Minor (optimization opportunity).
>
> If all indicated renoprotective therapies are already prescribed or contraindicated, state clearly: "No renoprotective therapy gaps identified" and explain why each class is either present or contraindicated.

Be thorough but precise. Only flag gaps where there is a clear guideline-supported indication AND no contraindication present in the patient data.

##### User Prompt Template

Please review the following CKD patient case and identify any renoprotective therapy omissions.

Patient Case:

`{case_json_formatted}`

Provide your complete assessment.

##### Notes

- This prompt is intentionally MORE focused than Study 01's open-ended CMR prompt
- The system prompt lists the 3 target classes explicitly — this tests whether focused guidance improves detection
- "Critical/Major/Minor" maps to Study 01's Level 3/2/1 for cross-study comparison
- JSON output is NOT required — free text allows richer clinical reasoning

ewpage

#### Supplementary Table S1: Multi-Gap Case Performance

Five cases in the cohort contain ≥2 prespecified gaps (gap-type combinations: G1+G2 SGLT2 inhibitor + finerenone). This analysis asks whether each LLM identified *all* gaps in such cases, not just one.

##### Summary

| Model | All gaps caught (n/30) | Partial | None | Per-gap recall |
|---|---|---|---|---|
| GPT-5.4 | 30 | 0 | 0 | 100.0% |
| Gemini 3.1 Pro | 30 | 0 | 0 | 100.0% |
| Grok 4.1 Fast | 30 | 0 | 0 | 100.0% |
| DeepSeek R1 | 29 | 1 | 0 | 98.3% |

##### Per-case detail

| Case | Gap types | GPT-5.4 | Gemini 3.1 Pro | Grok 4.1 Fast | DeepSeek R1 |
|---|---|---|---|---|---|
| GA-04 | SGLT2i+RAAS | 2/2 | 2/2 | 2/2 | 2/2 |
| GA-05 | SGLT2i+RAAS | 2/2 | 2/2 | 2/2 | 2/2 |
| GA-06 | SGLT2i+RAAS | 2/2 | 2/2 | 2/2 | 2/2 |
| GA-07 | SGLT2i+Finerenone | 2/2 | 2/2 | 2/2 | 2/2 |
| GA-08 | SGLT2i+Finerenone | 2/2 | 2/2 | 2/2 | 2/2 |
| GA-16 | SGLT2i+Finerenone | 2/2 | 2/2 | 2/2 | 1/2 |
| GA-17 | SGLT2i+Finerenone | 2/2 | 2/2 | 2/2 | 2/2 |
| GA-18 | SGLT2i+Finerenone | 2/2 | 2/2 | 2/2 | 2/2 |
| GA-19 | SGLT2i+Finerenone | 2/2 | 2/2 | 2/2 | 2/2 |
| GA-20 | SGLT2i+Finerenone | 2/2 | 2/2 | 2/2 | 2/2 |
| GA-21 | SGLT2i+Finerenone | 2/2 | 2/2 | 2/2 | 2/2 |
| GA-22 | SGLT2i+Finerenone | 2/2 | 2/2 | 2/2 | 2/2 |
| GA-23 | SGLT2i+Finerenone | 2/2 | 2/2 | 2/2 | 2/2 |
| GA-24 | SGLT2i+RAAS | 2/2 | 2/2 | 2/2 | 2/2 |
| GA-25 | SGLT2i+RAAS | 2/2 | 2/2 | 2/2 | 2/2 |

| Case | Gap types | GPT-5.4 | Gemini 3.1 Pro | Grok 4.1 Fast | DeepSeek R1 |
|---|---|---|---|---|---|
| GB-03 | SGLT2i+RAAS | 2/2 | 2/2 | 2/2 | 2/2 |
| GB-04 | SGLT2i+RAAS | 2/2 | 2/2 | 2/2 | 2/2 |
| GB-05 | SGLT2i+Finerenone | 2/2 | 2/2 | 2/2 | 2/2 |
| GB-06 | SGLT2i+Finerenone | 2/2 | 2/2 | 2/2 | 2/2 |
| GB-07 | SGLT2i+Finerenone | 2/2 | 2/2 | 2/2 | 2/2 |
| GB-16 | SGLT2i+Finerenone | 2/2 | 2/2 | 2/2 | 2/2 |
| GB-17 | SGLT2i+Finerenone | 2/2 | 2/2 | 2/2 | 2/2 |
| GB-18 | SGLT2i+Finerenone | 2/2 | 2/2 | 2/2 | 2/2 |
| GB-19 | SGLT2i+Finerenone | 2/2 | 2/2 | 2/2 | 2/2 |
| GB-20 | SGLT2i+Finerenone | 2/2 | 2/2 | 2/2 | 2/2 |
| GB-21 | SGLT2i+Finerenone | 2/2 | 2/2 | 2/2 | 2/2 |
| GB-22 | SGLT2i+Finerenone | 2/2 | 2/2 | 2/2 | 2/2 |
| GB-23 | SGLT2i+RAAS | 2/2 | 2/2 | 2/2 | 2/2 |
| GB-24 | SGLT2i+RAAS | 2/2 | 2/2 | 2/2 | 2/2 |
| GB-25 | SGLT2i+RAAS | 2/2 | 2/2 | 2/2 | 2/2 |

**Interpretation**

Models that identify the dominant gap (commonly SGLT2 inhibitor) but miss the secondary gap (commonly finerenone in G1+G2 multi-gap cases) would inflate apparent sensitivity if measured per case rather than per gap. Per-gap recall in the table above reflects the underlying performance correctly. Multi-gap performance is not a primary outcome but is reported to address the reviewer concern that high overall sensitivity could be driven by easy single-gap cases.

ewpage

### Supplementary Table S2: Clinically Incorrect Reasoning Analysis

**Definition.** Clinically incorrect reasoning is a factually incorrect or materially unsupported clinical claim made by the LLM that is not supported by the case data, current guidelines (KDIGO 2024), established pharmacology, or the study's prespecified operational criteria. Instances were identified during PI classification of each case and recorded with type and detail.

**Why this matters clinically.** Unlike sensitivity/specificity (which measure whether the LLM identified the correct gap), clinically incorrect reasoning rate measures whether the LLM's *reasoning text* contains misinformation that could mislead a clinician using the tool for decision support — even when the final recommendation happens to be correct.

**Summary by model**

| Model | Cases with clinically incorrect reasoning | Total instances | Rate (cases) | 95% Wilson CI |
|---|---:|---:|---:|---:|
| GPT-5.4 | 0/100 | 0 | 0.0% | [0.0%, 3.7%] |
| Gemini 3.1 Pro | 9/100 | 9 | 9.0% | [4.8%, 16.2%] |
| Grok 4.1 Fast | 10/100 | 10 | 10.0% | [5.5%, 17.4%] |
| DeepSeek R1 | 27/100 | 33 | 27.0% | [19.3%, 36.4%] |

**Clinically incorrect reasoning types by model**

| Type | GPT-5.4 | Gemini 3.1 Pro | Grok 4.1 Fast | DeepSeek R1 | Total |
|---|---:|---:|---:|---:|---:|
| fabricated_guideline | 0 | 2 | 0 | 1 | 3 |
| incorrect_pharmacology | 0 | 7 | 10 | 32 | 49 |

| Type | GPT-5.4 | Gemini 3.1 Pro | Grok 4.1 Fast | DeepSeek R1 | Total |
|---|---|---|---|---|---|

**Pairwise rate differences (descriptive; clinically incorrect reasoning rate)**

> **Note:** Clinically incorrect reasoning status is paired by case across models, violating the independence assumption of two-sample z-tests. Pairwise differences are therefore reported descriptively (rate difference + Wilson CIs per model) rather than with inferential p-values.

| Comparison | Rate difference |
|---|---|
| GPT-5.4 vs Gemini 3.1 Pro | -9.0 pp |
| GPT-5.4 vs Grok 4.1 Fast | -10.0 pp |
| GPT-5.4 vs DeepSeek R1 | -27.0 pp |
| Gemini 3.1 Pro vs Grok 4.1 Fast | -1.0 pp |
| Gemini 3.1 Pro vs DeepSeek R1 | -18.0 pp |
| Grok 4.1 Fast vs DeepSeek R1 | -17.0 pp |

**Full audit trail (all instances)**

| Model | Case | Type | Detail |
|---|---|---|---|
| Gemini 3.1 Pro | GB-03 | pharm (boundary) | LLM states RAAS blockade is typically deferred until potassium is <=5.0 mEq/L, whereas gold standard considers ACEi/ARB indicated at K 5.2 mEq/L (<=5.5). |
| Gemini 3.1 Pro | GB-12 | guideline (boundary) | LLM claims KDIGO explicitly recommends against initiating RAAS inhibitors when serum potassium is >5.0 mEq/L; this threshold is not supported by the gold standard, which considers ACEi/ARB indicated at potassium 5.4 mEq/L. |
| Gemini 3.1 Pro | GC-16 | pharm (boundary) | LLM states RAAS blockade initiation is clinically contraindicated at eGFR 10 and K 4.9, conflicting with the gold-standard KDIGO-based indication for ACEi/ARB in this case. |
| Gemini 3.1 Pro | GC-20 | guideline (factual) | LLM states guidelines advise against ACEi/ARB initiation if potassium >5.0 mEq/L, which does not match the gold-standard KDIGO-based criterion allowing initiation at K 5.1 mEq/L (<=5.5). |
| Gemini 3.1 Pro | GC-22 | pharm (boundary) | LLM states ACEi/ARB initiation is contraindicated due to eGFR 13 and potassium 5.2, which conflicts with the gold-standard KDIGO-based indication in this case. |
| Gemini 3.1 Pro | GC-24 | pharm (boundary) | LLM states ACEi/ARB initiation is clinically contraindicated solely because of CKD G5/eGFR 11 and borderline K 5.0, which is not the gold-standard/KDIGO-based interpretation for this case. |
| Gemini 3.1 Pro | GC-28 | pharm (boundary) | LLM states SGLT2 inhibitors are contraindicated for initiation in dialysis and eGFR <20; this overstates labeling/guideline restrictions as a blanket contraindication. |
| Gemini 3.1 Pro | GC-29 | pharm (boundary) | LLM states ACEi/ARB initiation is contraindicated solely due to eGFR 10 and potassium 4.9, which conflicts with the gold-standard KDIGO-based indication for RAAS blockade in this case. |
| Gemini 3.1 Pro | GC-30 | pharm (factual) | LLM states KDIGO 2024 does not recommend initiating ACEi/ARB in CKD G5 and characterizes initiation at eGFR 13/K 5.0 as unacceptably high risk; this conflicts with the gold-standard indication. |
| Grok 4.1 Fast | GB-03 | pharm (factual) | Claims losartan has lower hyperkalemia potential than ACE inhibitors, which is not a reliable class distinction. |

| Model | Case | Type | Detail |
| --- | --- | --- | --- |
| Grok 4.1 Fast | GB-04 | pharm (factual) | States ARB is preferred over ACEi in low eGFR to minimize initial hyperkalemia risk; this preference/rationale is not established. |
| Grok 4.1 Fast | GB-11 | pharm (factual) | LLM states ARB is preferred over ACEi in advanced CKD to minimize hyperkalemia risk; ARBs are not generally preferred over ACEi for this reason. |
| Grok 4.1 Fast | GB-25 | pharm (factual) | LLM states ARB is preferred over ACEi in CKD G4 due to lower hyperkalemia risk; this preference is not generally established as stated. |
| Grok 4.1 Fast | GC-16 | pharm (factual) | LLM states ARBs are preferred in elderly patients for lower risk of hyperkalemia vs ACE inhibitors; this is not correct as both ACEi and ARBs can raise potassium and ARBs are not generally preferred for lower hyperkalemia risk. |
| Grok 4.1 Fast | GC-20 | pharm (factual) | Claims ARB is preferred in advanced CKD for potentially lower hyperkalemia risk vs ACEi; this is not an established pharmacologic advantage. |
| Grok 4.1 Fast | GC-24 | pharm (factual) | Claimed ARB is preferred over ACEi due to lower hyperkalemia risk; this is not an established class advantage. |
| Grok 4.1 Fast | GC-28 | pharm (boundary) | LLM states RAAS blockade is contraindicated due to hyperkalemia at K 5.1 mEq/L and CKD G5D on HD, which conflicts with the gold standard indication threshold and overstates contraindication. |
| Grok 4.1 Fast | GC-29 | pharm (factual) | States ARB is preferred over ACEi in age >65 and low eGFR to minimize hyperkalemia risk; ARBs do not have a clear hyperkalemia advantage over ACE inhibitors. |
| Grok 4.1 Fast | GC-30 | pharm (factual) | LLM states ARB is preferred over ACEi in advanced CKD to minimize risk of acute worsening, which is not an established pharmacologic distinction. |
| DeepSeek R1 | GA-04 | pharm (factual) | Claimed empagliflozin has no sulfonamide moiety while dapagliflozin does; this distinction is inaccurate and not clinically meaningful. |
| DeepSeek R1 | GA-12 | pharm (factual) | States ARB preferred due to sulfonamide allergy; losartan is not selected based on sulfonamide cross-reactivity, and this rationale is pharmacologically unsound. |
| DeepSeek R1 | GA-24 | pharm (factual) | Incorrect claim that canagliflozin is a sulfa-containing SGLT2 inhibitor due to sulfonamide allergy concern. |
| DeepSeek R1 | GA-24 | pharm (factual) | Incorrect claim that irbesartan is a sulfa-containing ARB due to sulfonamide allergy concern. |
| DeepSeek R1 | GA-25 | pharm (factual) | States sodium bicarbonate may contribute to hyperkalemia; sodium bicarbonate is generally used to help mitigate hyperkalemia in CKD/metabolic acidosis, not cause it. |
| DeepSeek R1 | GB-02 | pharm (factual) | LLM states canagliflozin is an exception regarding sulfonamide content/cross-reactivity and prefers empagliflozin over dapagliflozin due to sulfonamide allergy; this is incorrect pharmacology. |
| DeepSeek R1 | GB-04 | pharm (factual) | Claims SGLT2 inhibitors contain a sulfonyl moiety and implies sulfonamide cross-reactivity concern. |
| DeepSeek R1 | GB-04 | pharm (factual) | States losartan avoids a sulfonamide moiety, which is not a relevant property of ACEi/ARB selection. |
| DeepSeek R1 | GB-09 | pharm (factual) | Finerenone dose adjustment/monitoring instructions are inaccurate; every-other-day reduction and 4-day potassium check are not standard labeled recommendations. |
| DeepSeek R1 | GB-10 | pharm (factual) | States ARB is preferred due to potassium 4.9 mEq/L; elevated potassium is not a reason to prefer ARB over ACEi, as both can raise potassium similarly. |

| Model | Case | Type | Detail |
|---|---|---|---|
| DeepSeek R1 | GB-18 | pharm (factual) | States both SGLT2 inhibitor and finerenone may increase potassium risk; SGLT2 inhibitors are not typically potassium-raising and may modestly lower hyperkalemia risk. |
| DeepSeek R1 | GB-22 | pharm (factual) | States dapagliflozin avoids a sulfa moiety and implies canagliflozin should be avoided for sulfonamide allergy; this is incorrect pharmacology. |
| DeepSeek R1 | GB-26 | guideline (factual) | Claimed KDIGO 2024 recommends avoiding SGLT2 inhibitor initiation if potassium >5.0 mEq/L. |
| DeepSeek R1 | GB-26 | pharm (factual) | Claimed SGLT2 inhibitors may exacerbate hyperkalemia; this is not correct pharmacology. |
| DeepSeek R1 | GB-33 | pharm (boundary) | States SGLT2 inhibitors are contraindicated below eGFR 20; this is not a general contraindication statement and is inaccurate framing. |
| DeepSeek R1 | GC-01 | pharm (boundary) | States ACEi/ARB is contraindicated/not routinely recommended solely due to CKD G5/eGFR 12 with K 4.8, which conflicts with KDIGO-based indication in this case. |
| DeepSeek R1 | GC-01 | pharm (boundary) | Calls SGLT2 inhibitor contraindicated because eGFR is below threshold; low eGFR here is a lack of indication/initiation threshold issue, not a true contraindication. |
| DeepSeek R1 | GC-01 | pharm (factual) | Calls finerenone contraindicated based on not meeting eligibility criteria; this is not equivalent to a pharmacologic contraindication. |
| DeepSeek R1 | GC-02 | pharm (factual) | Claimed sulfonamide allergy supports avoidance of SGLT2 inhibitors due to sulfonamide moieties. |
| DeepSeek R1 | GC-02 | pharm (factual) | Claimed sevelamer is a high-potassium medication contributing to hyperkalemia. |
| DeepSeek R1 | GC-04 | pharm (boundary) | LLM states KDIGO 2024 explicitly recommends discontinuing ACEi/ARB when patients transition to dialysis; this is incorrect for this case relative to the gold standard indication for ACEi/ARB. |
| DeepSeek R1 | GC-05 | pharm (boundary) | LLM states ACEi/ARB initiation is contraindicated due to K 5.4 and cites a threshold of >5.0 mEq/L, which conflicts with the gold-standard KDIGO-based interpretation for this case where K 5.4 still permits ACEi/ARB consideration. |
| DeepSeek R1 | GC-12 | pharm (factual) | Claimed sulfonamide allergy as a contraindication to SGLT2 inhibitors and stated SGLT2 inhibitors are sulfonamide derivatives. |
| DeepSeek R1 | GC-16 | pharm (boundary) | States ACEi/ARB are contraindicated/not routinely initiated in CKD G5 and lack proven renoprotection, which conflicts with the gold-standard KDIGO-based indication in this case. |
| DeepSeek R1 | GC-17 | pharm (boundary) | LLM states ACEi/ARB are contraindicated in CKD G5/eGFR 14 with K 5.0, which conflicts with the gold-standard indication for RAAS blockade here. |
| DeepSeek R1 | GC-18 | pharm (boundary) | LLM states ACEi/ARB are generally avoided/contraindicated in dialysis-dependent CKD and that KDIGO advises discontinuing or avoiding initiation in dialysis unless compelling indications exist; this conflicts with the gold standard for this case. |
| DeepSeek R1 | GC-19 | pharm (boundary) | LLM states ACEi/ARB are contraindicated for renoprotection solely because patient is dialysis-dependent/CKD G5D, contradicting the gold-standard indication used for this study case. |
| DeepSeek R1 | GC-20 | pharm (boundary) | LLM states potassium 5.1 mEq/L is above threshold for safe ACEi/ARB initiation and treats it as a contraindication, conflicting with the study gold standard threshold of <=5.5. |
| DeepSeek R1 | GC-22 | pharm (factual) | LLM states ACEi/ARB should be avoided if K >5.0 and labels K 5.2 as a contraindication, which is inconsistent with the study gold standard threshold allowing initiation at K <=5.5. |
| DeepSeek R1 | GC-24 | pharm (boundary) | LLM treated potassium 5.0 mEq/L as an absolute contraindication/barrier to ACEi/ARB initiation, conflicting with the case gold standard threshold. |

| Model | Case | Type | Detail |
|---|---|---|---|
| DeepSeek R1 | GC-25 | pharm (boundary) | LLM states ACEi/ARB are contraindicated because potassium 5.3 mEq/L is above target for initiation, conflicting with gold standard use of K <=5.5 as acceptable for initiation in this case. |
| DeepSeek R1 | GC-26 | pharm (factual) | Stated SGLT2 inhibitors require T2DM plus significant albuminuria or HFrEF; this is not correct KDIGO CKD indication logic. |
| DeepSeek R1 | GC-29 | pharm (boundary) | LLM states RAAS blockade may accelerate eGFR decline in stage G5 CKD and potentially precipitate dialysis; this is an incorrect pharmacology/generalization and contradicts guideline-supported ACEi/ARB use for albuminuric CKD unless specific contraindications exist. |

**Subtype breakdown:** 31 factual errors, 21 boundary-discordant conservative reasoning (total 52).

###### Clinical implications (for Discussion)

1. **Clinically incorrect reasoning rate dissociates from sensitivity.** A model may correctly identify the right therapy (high sensitivity) while embedding incorrect pharmacological reasoning (e.g., fabricated drug-class properties, wrong contraindication logic). For clinical-decision support, both signals matter: a clinician who trusts the reasoning text could be misled even when the final recommendation is correct.
2. **Inter-model differential is large.** The difference between the cleanest and noisiest model in this benchmark exceeds the difference in sensitivity, suggesting clinically incorrect reasoning rate is a more discriminating safety axis than detection accuracy alone.
3. **All factual errors identified are pharmacology-related** (drug class properties, dose interpretation, contraindication logic), which is the single most safety-critical category for renoprotective therapy decisions.

###### Methods note (for paper)

Clinically incorrect reasoning instances were operationalized as factually incorrect or materially unsupported clinical claims in the model's reasoning text, identified by the principal investigator during case classification. The PI judged each instance against KDIGO 2024 guidelines, current pharmacology references, and the case data; borderline statements were not counted. Rates are reported as case-level (proportion of cases containing >=1 clinically incorrect reasoning instance); instance counts are reported separately for granularity. Wilson 95% CIs are used for proportions; pairwise rate differences are reported descriptively (clinically incorrect reasoning status is paired by case, precluding standard two-sample tests). Instances are post-hoc stratified into factual errors vs boundary-discordant conservative reasoning.

ewpage

#### Supplementary Table S3: eGFR Threshold Robustness

**Why this analysis.** The primary subgroup analysis used eGFR<15 as the boundary cutoff. This sensitivity analysis tests whether the conservative-bias finding is robust to alternative threshold choices and whether the conservative behaviour activates at a sharp eGFR threshold or gradually as eGFR declines. A sharp inflection would support a *threshold-effect* interpretation (the model treats eGFR as a hard cutoff); a gradient would support a *graded conservatism* interpretation (every step lower in eGFR raises the bar incrementally).

###### RASi sensitivity at progressively wider eGFR cutoffs

| Stratum (RASi gap cases) | GPT-5.4 | Gemini 3.1 Pro | Grok 4.1 Fast | DeepSeek R1 |
|---|---|---|---|---|
| **eGFR < 10** (n=9) | 0/9 (0%) | 1/9 (11%) | 6/9 (67%) | 0/9 (0%) |
| **eGFR < 15 (primary boundary)** (n=20) | 11/20 (55%) | 2/20 (10%) | 17/20 (85%) | 2/20 (10%) |
| **eGFR < 20** (n=23) | 14/23 (61%) | 4/23 (17%) | 20/23 (87%) | 4/23 (17%) |
| **eGFR < 25** (n=27) | 18/27 (67%) | 8/27 (30%) | 24/27 (89%) | 8/27 (30%) |
| **eGFR ≥ 15 (non-boundary, reference)** (n=15) | 15/15 (100%) | 14/15 (93%) | 15/15 (100%) | 14/15 (93%) |
| **All RASi gap cases (overall)** (n=35) | 26/35 (74%) | 16/35 (46%) | 32/35 (91%) | 16/35 (46%) |

**Interpretation**

Reading top-to-bottom shows what happens to per-model sensitivity as the threshold widens from very advanced (eGFR<10) to all RASi gap cases. A model whose sensitivity rises sharply between two adjacent eGFR cuts has crossed an internal decision threshold; a model whose sensitivity rises smoothly has graded conservatism.

Practical clinical implications: 1. If the conservative bias activates already at eGFR<20 (broad boundary), the affected patient population is much larger than the eGFR<15 subgroup alone would suggest. 2. If sensitivity recovers fully only above eGFR≥30, the implication is that LLM-based decision support is least reliable across the entire G4–G5 range — exactly the patients in whom renoprotective therapy decisions are most consequential.

This analysis was added post-hoc as a robustness check and is reported as supplementary; it does not alter the prespecified primary boundary (eGFR<15).

ewpage

### Supplementary Table S4: Potassium Boundary Subgroup

**Why this analysis.** Hyperkalemia is the single most common real-world reason clinicians withhold or discontinue renoprotective therapy. KDIGO 2024 caps finerenone initiation at serum K ≤ 4.8; RASi is conventionally avoided when K > 5.5, with the 4.8–5.5 range being a clinical "yellow zone" in which different clinicians make different decisions. This analysis asks whether LLMs become more conservative as serum K approaches these thresholds — analogous to the eGFR<15 boundary finding in the primary analysis. Concordant patterns at two independent clinical boundaries (eGFR and K) would suggest a *generalisable* LLM tendency toward over-caution at decision boundaries, not a phenomenon specific to one variable.

##### Finerenone gap detection by serum potassium stratum

All finerenone gap cases by design satisfy K ≤ 4.8 (KDIGO eligibility); this analysis tests whether models miss finerenone gaps more often as K approaches that upper bound (4.6–4.8) than in clearly safe cases (K ≤ 4.5).

| Stratum (finerenone gap cases) | GPT-5.4 | Gemini 3.1 Pro | Grok 4.1 Fast | DeepSeek R1 |
|---|---|---|---|---|
| **K ≤ 4.5** (n=23) | 23/23 (100%) | 23/23 (100%) | 23/23 (100%) | 22/23 (96%) |
| **K 4.6 – 4.8 (KDIGO upper bound)** (n=11) | 11/11 (100%) | 11/11 (100%) | 11/11 (100%) | 11/11 (100%) |
| **All finerenone gap cases** (n=34) | 34/34 (100%) | 34/34 (100%) | 34/34 (100%) | 33/34 (97%) |

**RASi gap detection by serum potassium stratum**

All RASi gap cases by design satisfy K ≤ 5.5 (RASi eligibility threshold); this analysis tests whether models miss RASi gaps more often as K rises into the 4.9–5.5 "yellow zone" than in clearly safe cases (K ≤ 4.8).

| Stratum (RASi gap cases) | GPT-5.4 | Gemini 3.1 Pro | Grok 4.1 Fast | DeepSeek R1 |
|---|---|---|---|---|
| **K ≤ 4.8** (n=17) | 11/17 (65%) | 11/17 (65%) | 16/17 (94%) | 10/17 (59%) |
| **K 4.9 – 5.0** (n=11) | 10/11 (91%) | 4/11 (36%) | 11/11 (100%) | 5/11 (45%) |
| **K 5.1 – 5.5 (yellow zone)** (n=7) | 5/7 (71%) | 1/7 (14%) | 5/7 (71%) | 1/7 (14%) |
| **All RASi gap cases** (n=35) | 26/35 (74%) | 16/35 (46%) | 32/35 (91%) | 16/35 (46%) |

**Interpretation**

**Finerenone:** A drop in sensitivity in the K 4.6–4.8 stratum compared with K ≤ 4.5 would indicate that models are reluctant to recommend finerenone when the patient is already at the KDIGO-allowed upper potassium bound — even though the eligibility criterion is technically met. This would be a clinically meaningful miss because patients at the borderline are precisely those whose future K trajectory matters most.

**RASi:** A drop in sensitivity as K moves into the 4.9–5.5 yellow zone would indicate models reproduce real-world physician behaviour of withholding RASi at K levels that are still within current eligibility. If the same conservative-bias regime activates at both eGFR<15 (primary) and K 4.9–5.5 (secondary), the implication is a *generalisable boundary-aversion phenomenon* in current LLMs — relevant to any clinical-decision-support deployment in which a drug has a lab-defined safety threshold.

This analysis was added post-hoc as a robustness check and extension; pairwise statistical comparisons within strata are not reported because the n per stratum is modest. Effect magnitudes are interpreted descriptively.

ewpage

### Supplementary Table S5: Replicate-Query Sensitivity

Sensitivity computed at the gap level (n = 105 prespecified gaps). Run 1 = single-query deployment from the first extracted run; Consensus = PI-scored consensus-selected decision used in the primary analysis; Worst-case = gap detected only if ALL 3 runs detected it; Best-case = gap detected if ANY run detected it.

| Model | Run 1 | Consensus | Worst-case | Best-case |
|---|---|---|---|---|
| GPT-5.4 | 91.4% (96/105) | 91.4% (96/105) | 90.5% (95/105) | 92.4% (97/105) |
| Gemini 3.1 Pro | 85.7% (90/105) | 81.9% (86/105) | 81.9% (86/105) | 88.6% (93/105) |
| Grok 4.1 Fast | 97.1% (102/105) | 97.1% (102/105) | 96.2% (101/105) | 98.1% (103/105) |
| DeepSeek R1 | 83.8% (88/105) | 80.0% (84/105) | 74.3% (78/105) | 89.5% (94/105) |

ewpage

#### Supplementary Table S6: Pairwise McNemar Tests

Per-gap-type pairwise model comparisons. The omnibus Cochran's Q tests for SGLT2 inhibitor and finerenone omissions were non-significant (Q=3.00, p=0.39 for both), so the pairwise tables for these gap types are reported here for completeness only. The substantive between-model differences are confined to the RASi (ACEi/ARB) comparisons.

##### Pairwise McNemar Tests with Holm-Bonferroni Correction

Paired binary comparison (gap detected = 1, missed = 0) per case. b/c are discordant pairs (b: m1 missed, m2 caught; c: m1 caught, m2 missed). Exact binomial used when discordant pairs <25.

**SGLT2i (n=36) -** ***Omnibus Cochran's Q non-significant (Q=3.00, p=0.39); pairwise tests are descriptive only***

| Comparison | b | c | statistic | method | p (raw) | p (Holm) | Sig. (Holm α=0.05) |
|---|---|---|---|---|---|---|---|
| GPT-5.4 vs Gemini 3.1 Pro | 0 | 0 | 0.000 | no discordant pairs | 1.0000 | 1.0000 | No |
| GPT-5.4 vs Grok 4.1 Fast | 0 | 0 | 0.000 | no discordant pairs | 1.0000 | 1.0000 | No |
| GPT-5.4 vs DeepSeek R1 | 0 | 1 | 0.000 | exact binomial | 1.0000 | 1.0000 | No |
| Gemini 3.1 Pro vs Grok 4.1 Fast | 0 | 0 | 0.000 | no discordant pairs | 1.0000 | 1.0000 | No |
| Gemini 3.1 Pro vs DeepSeek R1 | 0 | 1 | 0.000 | exact binomial | 1.0000 | 1.0000 | No |
| Grok 4.1 Fast vs DeepSeek R1 | 0 | 1 | 0.000 | exact binomial | 1.0000 | 1.0000 | No |

**Finerenone (n=34) -** ***Omnibus Cochran's Q non-significant (Q=3.00, p=0.39); pairwise tests are descriptive only***

| Comparison | b | c | statistic | method | p (raw) | p (Holm) | Sig. (Holm α=0.05) |
|---|---|---|---|---|---|---|---|
| GPT-5.4 vs Gemini 3.1 Pro | 0 | 0 | 0.000 | no discordant pairs | 1.0000 | 1.0000 | No |
| GPT-5.4 vs Grok 4.1 Fast | 0 | 0 | 0.000 | no discordant pairs | 1.0000 | 1.0000 | No |

| Comparison | b | c | statistic | method | p (raw) | p (Holm) | Sig. (Holm α=0.05) |
|---|---|---|---|---|---|---|---|
| GPT-5.4 vs DeepSeek R1 | 0 | 1 | 0.000 | exact binomial | 1.0000 | 1.0000 | No |
| Gemini 3.1 Pro vs Grok 4.1 Fast | 0 | 0 | 0.000 | no discor-dant pairs | 1.0000 | 1.0000 | No |
| Gemini 3.1 Pro vs DeepSeek R1 | 0 | 1 | 0.000 | exact binomial | 1.0000 | 1.0000 | No |
| Grok 4.1 Fast vs DeepSeek R1 | 0 | 1 | 0.000 | exact binomial | 1.0000 | 1.0000 | No |

**RASi (ACEi/ARB) (n=35)**

| Comparison | b | c | statistic | method | p (raw) | p (Holm) | Sig. (Holm α=0.05) |
|---|---|---|---|---|---|---|---|
| GPT-5.4 vs Gemini 3.1 Pro | 1 | 11 | 1.000 | exact binomial | 0.0063 | 0.0190 | **Yes** |
| GPT-5.4 vs Grok 4.1 Fast | 6 | 0 | 0.000 | exact binomial | 0.0312 | 0.0625 | No |
| GPT-5.4 vs DeepSeek R1 | 0 | 10 | 0.000 | exact binomial | 0.0020 | 0.0078 | **Yes** |
| Gemini 3.1 Pro vs Grok 4.1 Fast | 16 | 0 | 0.000 | exact binomial | 0.0000 | 0.0002 | **Yes** |
| Gemini 3.1 Pro vs DeepSeek R1 | 1 | 1 | 1.000 | exact binomial | 1.0000 | 1.0000 | No |
| Grok 4.1 Fast vs DeepSeek R1 | 0 | 16 | 0.000 | exact binomial | 0.0000 | 0.0002 | **Yes** |
